# The Survey of the Health of Wisconsin (SHOW) Program: An infrastructure for Advancing Population Health Sciences

**DOI:** 10.1101/2021.03.15.21253478

**Authors:** Kristen M.C. Malecki, Maria Nikodemova, Amy A. Schultz, Tamara J. LeCaire, Andrew J. Bersch, Lisa Cadmus-Bertram, Corinne D. Engelman, Erika Hagen, Mari Palta, Ajay K. Sethi, Matt C. Walsh, F. Javier Nieto, Paul E. Peppard

## Abstract

**Purpose:** The Survey of the Health of Wisconsin (SHOW) was established in 2008 by the University of Wisconsin (UW) School of Medicine and Public Health (SMPH) with the goals of 1) providing a timely and accurate picture of the health of the state residents; and 2) serving as an agile resource infrastructure for ancillary studies. Today SHOW continues to serve as a vital population health research infrastructure.

**Participants:** SHOW currently includes 5,846 adult and 980 minor participants recruited between 2008-2019 in four primary waves. WAVE I (2008-2013) includes annual statewide representative samples of 3,380 adults ages 21 to 74 years. WAVE II (2014-2016) is a triannual statewide sample of 1957 adults (age ≥18 years) and 645 children. WAVE III (2017) consists of follow-up of 725 adults from the WAVE I and baseline surveys of 222 children in selected households. WAVEs II and III include stool samples collected as part of an ancillary study in a subset of 784 individuals. WAVE IV consist of 517 adults and 113 children recruited from traditionally under-represented populations in biomedical research including African Americans and Hispanics in Milwaukee county, WI.

**Findings to Date:** The SHOW provides extensive data to examine the intersectionality of multiple social determinants and population health. SHOW includes a large biorepository and extensive health data collected in a geographically diverse urban and rural population. Over 60 studies have been published covering a broad range of topics including, urban and rural disparities in cardio-metabolic disease and cancer, objective physical activity, sleep, green-space and mental health, transcriptomics, the gut microbiome, antibiotic resistance, air pollution, concentrated animal feeding operations and heavy metal exposures.

**Future Plans:** The SHOW cohort is available for continued longitudinal follow-up and ancillary studies including genetic, multi-omic and translational environmental health, aging, microbiome and COVID-19 research.

**Article Summary:** *Strengths and limitations:*

- The Survey of the Health of Wisconsin (SHOW) is an infrastructure to advance population health sciences including biological sample collection and broader data on individual and neighborhood social and environmental determinants of health.
- The extensive data from diverse urban and rural populations offers a unique study sample to compare how socio-economic gradients shape health outcomes in different contexts.
- The objective health data supports novel interdisciplinary research initiatives and is especially suited for research in causes and consequences of environmental exposures (physical, chemical, social) across the life course on cardiometabolic health, immunity, and aging related conditions.
- The extensive biorepository supports novel omics research into common biological mechanisms underlying numerous complex chronic conditions including inflammation, oxidative stress, metabolomics, and epigenetic modulation.
- Ancillary studies, such as the Wisconsin Microbiome Study, have expanded the utility of the study to examine human susceptibility to environmental exposures and opportunities for investigations of the role of microbiome in health and disease.
- Long-standing partnerships and recent participation among traditionally under-represented populations in biomedical research offer numerous opportunities to support community-driven health equity work.
- No biological samples were collected among children.
- The statewide sampling frame may limit generalizability to other regions in the United States.

## Introduction

### Why was the cohort set up?

Increasingly, it is understood that health and well-being are shaped by a myriad of interconnected factors. These factors operate at multiple levels from individual differences in genetics, environmental exposures and life experiences to the physical environment, social, cultural and economic contexts in which we live. Several long-term general population cohort studies, such as the Framingham Heart Study the Nurse’s Health Study that began in mid-20^th^ century,^1-3^ have provided extensive information on determinants of priority health conditions including cardiovascular disease and cancer in the United States. Technological advances are leading to rapid discovery of biological markers and new population-based research infrastructures are needed to support novel translational research. Further, new resources are needed to capture the multi-level data with increased geographic granularity are necessary to advance population health sciences. Next-generation sequencing and “big-data” approaches have accelerated the pace of biomarker discovery, but how these biological factors are shaped by larger social, environmental and individual-level determinants within and between diverse populations across the lifecourse is less well understood.

The Survey of the Health of Wisconsin (SHOW) was established in 2008 by the University of Wisconsin School of Medicine and Public Health with funding from an institutional endowment with the goals of 1) providing a timely and accurate picture of the health of the state residents; and 2) serving as an agile resource infrastructure for ancillary studies that require access to community-based samples. Initially modeled after National Health and Nutrition Examination Survey (NHANES), SHOW provides a level of granularity to study the health status of individuals and determinants across rural and urban areas at a greater level of detail than national surveys.^4^ A decade later, the SHOW study sample continues to grow through multiple waves of data collection and ancillary studies and continues to serve as a state-of-the-art infrastructure for population health research. SHOWs mission in 2021 is to ***support ongoing population health monitoring and research, foster diverse partnerships, and support ongoing education in order to promote population health equity and well-being*** in Wisconsin and beyond. Core funding for SHOW is provided by the Wisconsin Partnership Program and additional support comes from ancillary projects funded by the National Institutes of Health and the Wisconsin Department of Health Services, among others. Scientific direction is provided by experts in population health research from across the entire University of Wisconsin-Madison campus, including a Scientific Advisory Board.

Unique elements of the SHOW program include the geographically diverse study population, the breadth of objective and biological data collected, the ability to link social and environmental contextual data, and the flexibility of the program to support translational science and health equity research. To date, no other such statewide study sample exists. From its inception, SHOW aimed to capture multi-level determinants of data to examine proximate and distal factors shaping health and well-being. Questionnaire data include a variety of medical and family history, mental health, occupation, life experiences, physical activity, diet, sleep and neighborhood perception data. The detailed data on household address and residential history can now be integrated with objective health and biomarker data to support innovative research projects integrating contextual social and environmental data across the life-course with cutting-edge biomarker analyses to advance understanding of biological mechanisms underlying health inequalities. Field data collection continues today with numerous opportunities for investigators to inform longitudinal follow-up and clinical collaborations including opportunities for linkage with electronic health records and other administrative data.

SHOW provides a unique resource to advance population health and aging research. A substantial portion of the SHOW study sample are middle to older adults born during the last quarter of the 20^th^ or early 21^st^ century. Many lifestyle, economic and social factors have changed within these generations and SHOW offers numerous opportunities to examine how these complex factors now influence health and well-being as participants age. More recently, focused recruitment efforts have aimed to expand the core study population to include children and increase the racial, ethnic and socio-economic diversity of the study population.

### Cohort Description

The full study sample includes 5,846 adult (ages 18 years and over) and 980 minor (age 0-17 years) participants. **Table 1** depicts the multiple waves of data collection and highlights key additions and changes to the cohort composition, sampling strategy, inclusion and exclusion criteria and study components over time. In brief, participants have been recruited across three waves (WAVE 1: 2008-2013, WAVE II: 2014-2016, and WAVE IV: 2018-2019). The first follow-up of WAVE I participants was completed in 2017, and is referred to as WAVE III. Ongoing retention of the SHOW cohort is maintained through community outreach, dissemination of study findings online and at community events. Bi-annual newsletters are also disseminated via mail and email to facilitate successful follow-up. Standard SHOW protocols are implemented consistently across each wave of data and biosample collection. All methods are well-documented through meta-data and online codebooks to ensure rigor and reproducibility over time. Supplemental Table 1A shows improvement in response rates, measured as number of participants screened eligible willing to participate in the program, over time, by health region and ten counties corresponding to each health region. Health regions are defined as geographic clusters of counties within a public health service area defined by the Wisconsin Department of Health Services. Supplemental Table 1B shows response rates by urbanicity as defined by the U.S. Census. Details regarding the design and data collection for each wave of recruitment and data collection for the SHOW study to date are briefly described below.

**Table 1.**
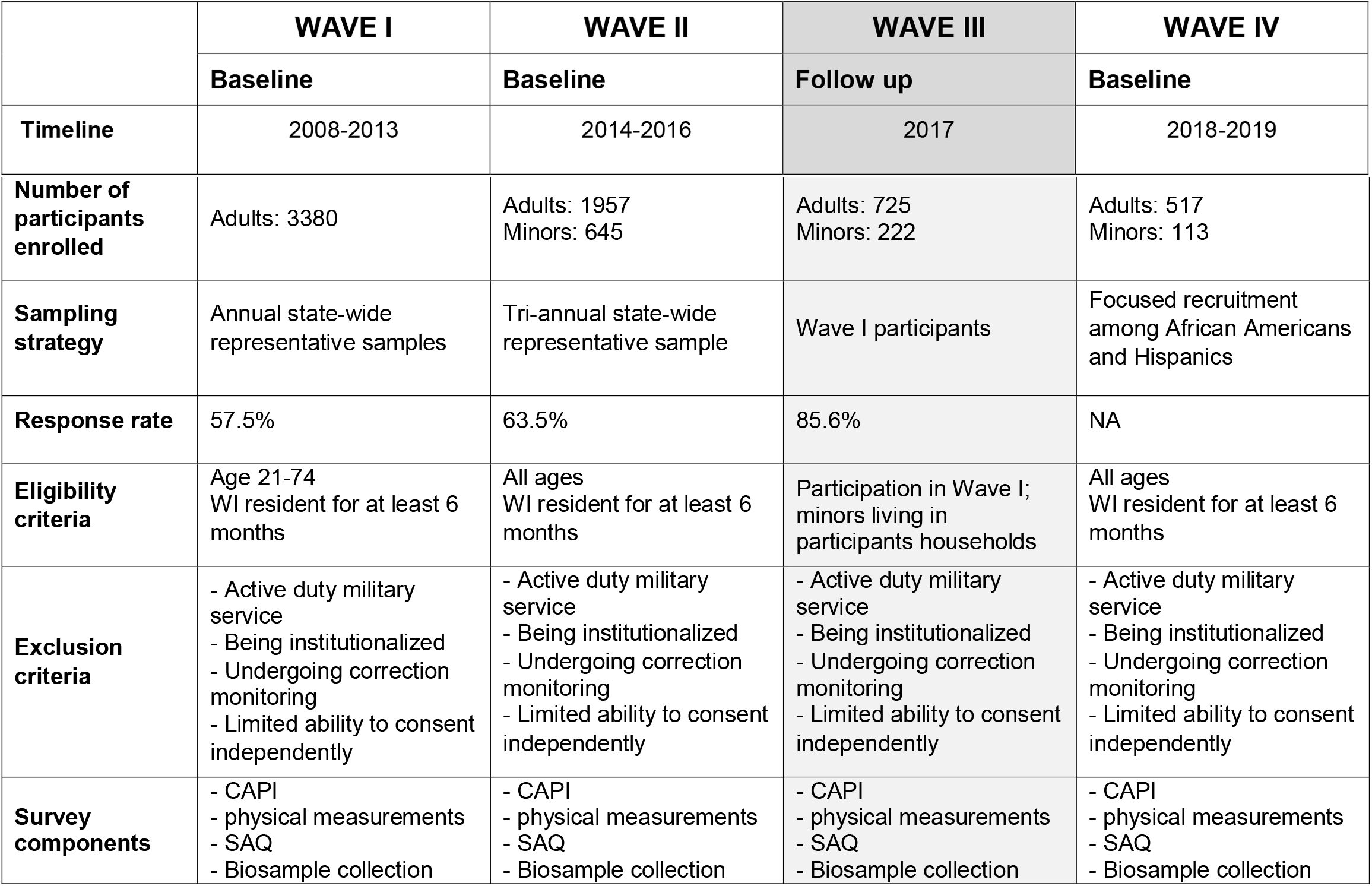
SHOW Survey Participant Summary, Sampling Strategy and Components by WAVE.

|  | WAVE I | WAVE II | WAVE III | WAVE IV |
| --- | --- | --- | --- | --- |
|  | Baseline | Baseline | Follow up | Baseline |
| Timeline | 2008-2013 | 2014-2016 | 2017 | 2018-2019 |
| **Number of participants enrolled** | Adults: 3380 | Adults: 1957<br>Minors: 645 | Adults: 725<br>Minors: 222 | Adults: 517<br>Minors: 113 |
| **Sampling strategy** | Annual state-wide representative samples | Tri-annual state-wide representative sample | Wave I participants | Focused recruitment among African Americans and Hispanics |
| **Response rate** | 57.5% | 63.5% | 85.6% | NA |
| **Eligibility criteria** | Age 21-74<br>WI resident for at least 6 months | All ages<br>WI resident for at least 6 months | Participation in Wave I; minors living in participants households | All ages<br>WI resident for at least 6 months |
| **Exclusion criteria** | - Active duty military service<br>- Being institutionalized<br>- Undergoing correction monitoring<br>- Limited ability to consent independently | - Active duty military service<br>- Being institutionalized<br>- Undergoing correction monitoring<br>- Limited ability to consent independently | - Active duty military service<br>- Being institutionalized<br>- Undergoing correction monitoring<br>- Limited ability to consent independently | - Active duty military service<br>- Being institutionalized<br>- Undergoing correction monitoring<br>- Limited ability to consent independently |
| **Survey components** | - CAPI<br>- physical measurements<br>- SAQ<br>- Biosample collection | - CAPI<br>- physical measurements<br>- SAQ<br>- Biosample collection | - CAPI<br>- physical measurements<br>- SAQ<br>- Biosample collection | - CAPI<br>- physical measurements<br>- SAQ<br>- Biosample collection |

### WAVE I - The Original SHOW Study Sample (2008-2013)

WAVE I (2008-2013) includes annual statewide representative samples of 3,380 adults ages 21 to 74 years with key demographics presented in Table 1A. As previously described by Nieto et al., 2010, a state-wide address-based sampling frame and two-stage, area probability sampling without replacement (PPSWOR) was used to generate an annual statewide representative sample.^56^ Selection criteria included age between 21-74 years, and residency within the state for longer than six months. Exclusion criteria included limited ability to consent independently, active-duty military service, being institutionalized, and undergoing community or home corrections monitoring. The annual sample size ranged from approximately 300 to 900 between 2008 and 2013. Response rates ranged from 43-87% depending on region across the state and, on average, tended to be higher in rural communities and lower in urban and lower income communities (Supplemental Table 1). Approximately 80% of participants who completed the household interview went on to complete all survey components (personal in-home interviews, self-administered questionnaire, physical exam, and biosample collection). Survey weights that incorporate design weights and adjustments for non-response and post-stratification, calibrated to the U.S. Census 2010 population totals by age, sex and race, improve the representativeness of statewide estimates, and design variables account for spatial clustering in the sample design.

### WAVE II - SHOW Expansion (2014-2016)

WAVE II, SHOW 2014-2016, provided a newly recruited prospective tri-annual statewide representative sample of 1957 adults (age ≥18 years) and 645 children (<18 years of age). Demographic data for the adult sample are presented in Table 1A while children are presented in Table 1C. Eligibility criteria for WAVE II expanded to add children (<18) and adults over age 74 years. Exclusion criteria were the same as for Wave I. Similar to WAVE I, an area probability sampling design was used to randomly select households, where all eligible household members were invited to participate. The main change between the waves was that the two-stage sampling design was modified to three-stages with county as the primary sampling unit (PSU) rather than Census block group (CBG). Eight PSUs, stratified by years of potential life lost, were randomly selected with probabilities proportional to size where the measure of size was occupied housing units. Two counties (Milwaukee and Dane) were selected with certainty (probability of selection=1) based on their large number of occupied housing units relative to the other counties. CBGs served as secondary sampling units with poverty stratification, and households within each CBG were randomly selected using simple random sampling.

Response rates were slightly higher on average in WAVE II with 64% of screened eligible individuals agreeing to participate. This higher response rate was attributed to additional focus on identifying field interviewers representative of the targeted community, and additional focus on community engagement and awareness campaigns, including endorsement by local officials prior to recruitment. Finally, we aimed to improve the ease of exam visits and sample collection by identifying exam visit locations in places of worship, or other locally respected locations that were convenient and centrally located for study participants. Design variables that account for clustering in the sampling design and survey weights based on design weights adjusted for non-response and calibrated to the U.S. Census Current Population Survey 2016 estimates by age, sex and race are available for WAVE II.

### WAVE III – Follow-up

WAVE III included longitudinal follow-up of n=725 adults from WAVE I (see Table 1B) and baseline participation of 222 children (see Table 1C). The eligibility criteria for WAVE III were participation in WAVE I, consent to be contacted by SHOW for future studies, WAVE I residence in 13 select counties cover the full spectrum of urbanicity and county health rankings across Wisconsin. For Non-Hispanic white participants, additional eligibility criteria were completion of the physical examination and biomarker collection in Wave I. All children currently residing in follow-up participant households were also eligible.

WAVE III follow-up included an in-home interview, physical exam, core biospecimen collection (blood, urine) and stool and skin swabs collection for microbiome analysis funded via ancillary study funding described below. Follow-up participation rate, determined based on number of those contacted who agreed to participate again, was estimated at 86% (see Supplemental Figure 1). Survey weights were not generated for WAVE III since it was not a random subsample of WAVE I.

### WAVE IV – Focused Recruitment of Traditionally Under-Represented Populations in Biomedical Research

In 2018-2019 SHOW focused on engaging and recruiting participants from two traditionally under-represented populations in biomedical research including an oversample of 440 African American (339 adult and 101 minor) and 131 Hispanic (125 adult and 6 minor-See Table 2C) participants living in and around the City of Milwaukee (see Table 2B for demographic details on adults). Unlike in WAVES I and II, both two-stage area probability sampling and community engaged convenience sampling approaches using community-based events were employed as primary recruitment strategies. The two-stage area probability sampling design was analogous to WAVE I, with the exception that the PSU sampling frame was restricted to 236 CBGs in the City of Milwaukee with populations of at least 60% African Americans based on the American Community Survey from 2015.

**Table 2-A.**
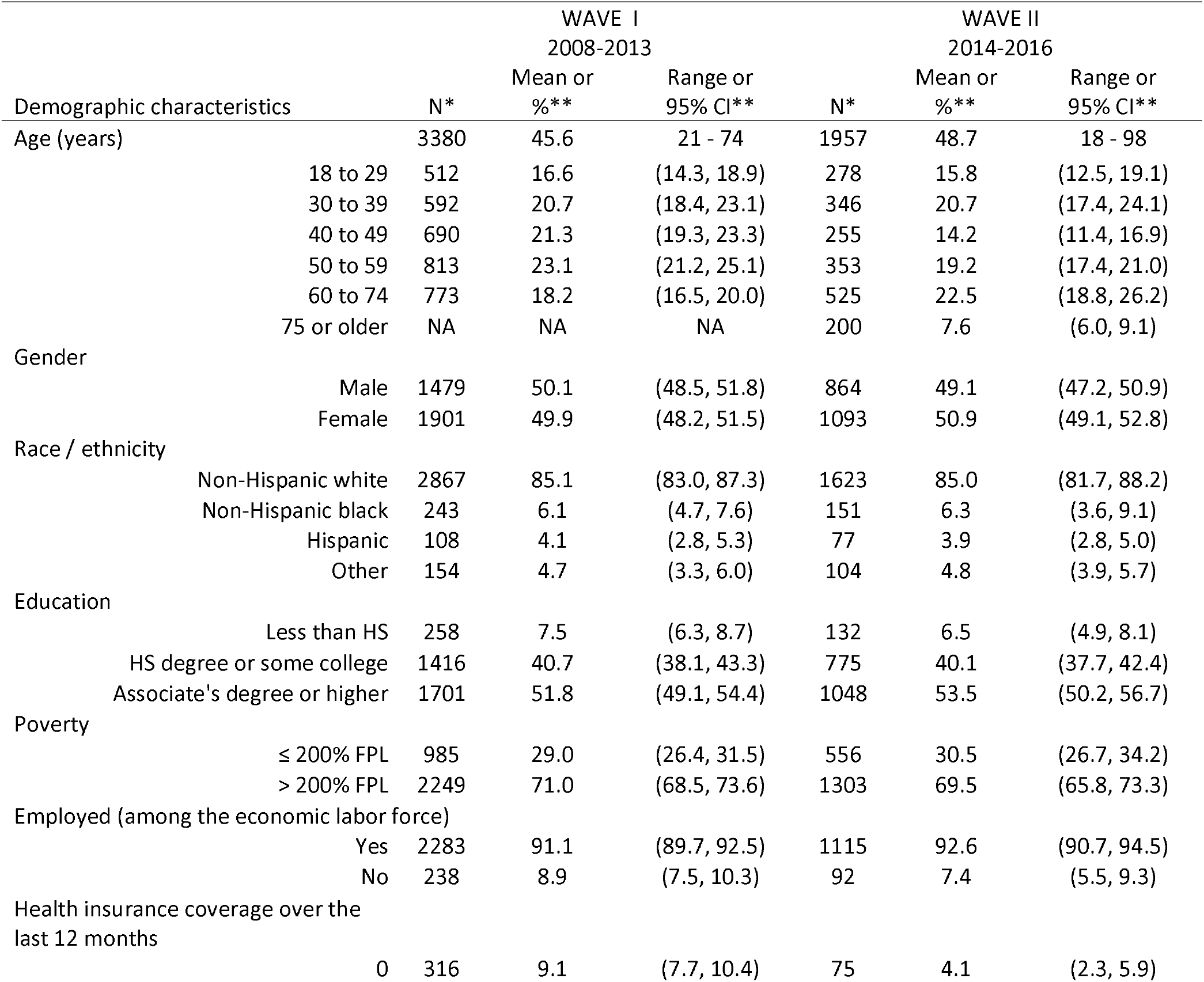
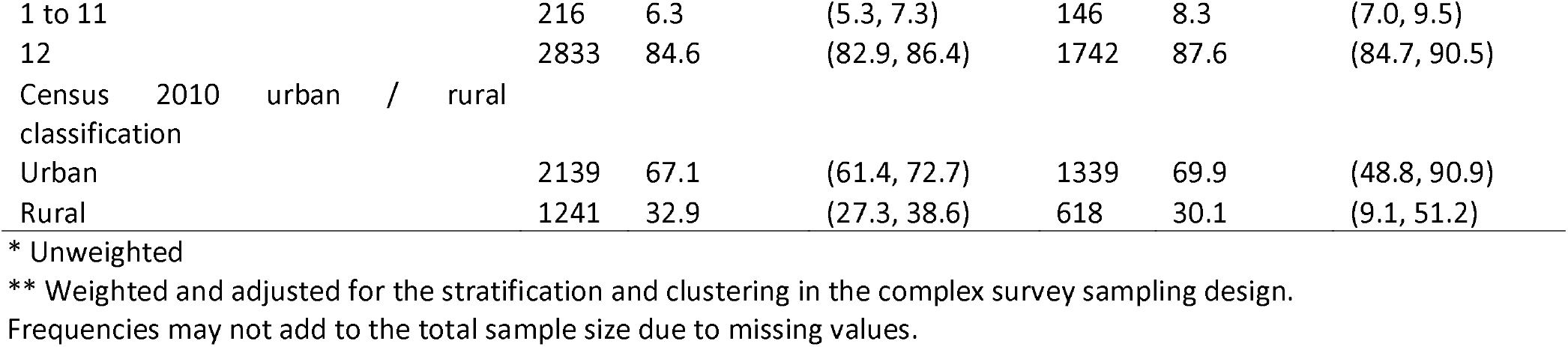
SHOW Adults WAVES I and II Characteristics, Weighted for Statewide Sample Estimation.

|  |  | WAVE I<br>2008-2013 |  |  | WAVE II<br>2014-2016 |  |
| --- | --- | --- | --- | --- | --- | --- |
| Demographic characteristics | N* | Mean or<br>%** | Range or<br>95% CI** | N* | Mean or<br>%** | Range or<br>95% CI** |
| Age (years) | 3380 | 45.6 | 21 - 74 | 1957 | 48.7 | 18 - 98 |
| 18 to 29 | 512 | 16.6 | (14.3, 18.9) | 278 | 15.8 | (12.5, 19.1) |
| 30 to 39 | 592 | 20.7 | (18.4, 23.1) | 346 | 20.7 | (17.4, 24.1) |
| 40 to 49 | 690 | 21.3 | (19.3, 23.3) | 255 | 14.2 | (11.4, 16.9) |
| 50 to 59 | 813 | 23.1 | (21.2, 25.1) | 353 | 19.2 | (17.4, 21.0) |
| 60 to 74 | 773 | 18.2 | (16.5, 20.0) | 525 | 22.5 | (18.8, 26.2) |
| 75 or older | NA | NA | NA | 200 | 7.6 | (6.0, 9.1) |
| Gender |  |  |  |  |  |  |
| Male | 1479 | 50.1 | (48.5, 51.8) | 864 | 49.1 | (47.2, 50.9) |
| Female | 1901 | 49.9 | (48.2, 51.5) | 1093 | 50.9 | (49.1, 52.8) |
| Race / ethnicity |  |  |  |  |  |  |
| Non-Hispanic white | 2867 | 85.1 | (83.0, 87.3) | 1623 | 85.0 | (81.7, 88.2) |
| Non-Hispanic black | 243 | 6.1 | (4.7, 7.6) | 151 | 6.3 | (3.6, 9.1) |
| Hispanic | 108 | 4.1 | (2.8, 5.3) | 77 | 3.9 | (2.8, 5.0) |
| Other | 154 | 4.7 | (3.3, 6.0) | 104 | 4.8 | (3.9, 5.7) |
| Education |  |  |  |  |  |  |
| Less than HS | 258 | 7.5 | (6.3, 8.7) | 132 | 6.5 | (4.9, 8.1) |
| HS degree or some college | 1416 | 40.7 | (38.1, 43.3) | 775 | 40.1 | (37.7, 42.4) |
| Associate's degree or higher | 1701 | 51.8 | (49.1, 54.4) | 1048 | 53.5 | (50.2, 56.7) |
| Poverty |  |  |  |  |  |  |
| ≤ 200% FPL | 985 | 29.0 | (26.4, 31.5) | 556 | 30.5 | (26.7, 34.2) |
| > 200% FPL | 2249 | 71.0 | (68.5, 73.6) | 1303 | 69.5 | (65.8, 73.3) |
| Employed (among the economic labor force) |  |  |  |  |  |  |
| Yes | 2283 | 91.1 | (89.7, 92.5) | 1115 | 92.6 | (90.7, 94.5) |
| No | 238 | 8.9 | (7.5, 10.3) | 92 | 7.4 | (5.5, 9.3) |
| Health insurance coverage over the<br>last 12 months |  |  |  |  |  |  |
| 0 | 316 | 9.1 | (7.7, 10.4) | 75 | 4.1 | (2.3, 5.9) |
| 1 to 11 | 216 | 6.3 | (5.3, 7.3) | 146 | 8.3 | (7.0, 9.5) |
| 12 | 2833 | 84.6 | (82.9, 86.4) | 1742 | 87.6 | (84.7, 90.5) |
| Census 2010 urban / rural classification |  |  |  |  |  |  |
| Urban | 2139 | 67.1 | (61.4, 72.7) | 1339 | 69.9 | (48.8, 90.9) |
| Rural | 1241 | 32.9 | (27.3, 38.6) | 618 | 30.1 | (9.1, 51.2) |

\* Unweighted
\*\* Weighted and adjusted for the stratification and clustering in the complex survey sampling design.
Frequencies may not add to the total sample size due to missing values.

**Table 2-B.** SHOW Adults WAVES III and IV Characteristics, Unweighted.

| Demographic characteristics | WAVE III<br>Follow-up 2017 |  |  | WAVE IV<br>Focused Population Oversample |  |  |
| --- | --- | --- | --- | --- | --- | --- |
|  | N | Mean or % | Range or 95% CI | N | Mean or % | Range or 95% CI |
| Age (years) | 725 | 54.1 | 25 - 82 | 517 | 46.8 | 18 - 91 |
| 18 to 29 | 29 | 4.0 | (2.6, 5.4) | 92 | 17.8 | (14.5, 21.1) |
| 30 to 39 | 114 | 15.7 | (13.1, 18.4) | 94 | 18.2 | (14.8, 21.5) |
| 40 to 49 | 128 | 17.7 | (14.9, 20.4) | 94 | 18.2 | (14.8, 21.5) |
| 50 to 59 | 157 | 21.7 | (18.6, 24.7) | 110 | 21.3 | (17.7, 24.8) |
| 60 to 74 | 238 | 32.8 | (29.4, 36.3) | 111 | 21.5 | (17.9, 25.0) |
| 75 or older | 59 | 8.1 | (6.1, 10.1) | 16 | 3.1 | (1.6, 4.6) |
| Gender |  |  |  |  |  |  |
| Male | 288 | 39.7 | (36.2, 43.3) | 199 | 38.5 | (34.3, 42.7) |
| Female | 437 | 60.3 | (56.7, 63.8) | 318 | 61.5 | (57.3, 65.7) |
| Race / ethnicity |  |  |  |  |  |  |
| Non-Hispanic white | 575 | 79.5 | (76.6, 82.5) | 33 | 6.4 | (4.3, 8.5) |
| Non-Hispanic black | 96 | 13.3 | (10.8, 15.8) | 339 | 65.6 | (61.5, 69.7) |
| Hispanic | 22 | 3.0 | (1.8, 4.3) | 125 | 24.2 | (20.5, 27.9) |
| Other | 30 | 4.1 | (2.7, 5.6) | 20 | 3.9 | (2.2, 5.5) |
| Education |  |  |  |  |  |  |
| Less than HS | 47 | 6.5 | (4.7, 8.3) | 159 | 30.8 | (26.8, 34.7) |
| HS degree or some college | 272 | 37.5 | (34.0, 41.0) | 249 | 48.2 | (43.8, 52.5) |
| Associate's degree or higher | 406 | 56.0 | (52.4, 59.6) | 109 | 21.1 | (17.6, 24.6) |
| Poverty |  |  |  |  |  |  |
| ≤ 200% FPL | 167 | 23.7 | (20.5, 26.8) | 344 | 74.9 | (71.0, 78.9) |
| > 200% FPL | 539 | 76.3 | (72.2, 79.5) | 115 | 25.1 | (21.1, 29.0) |
| Employed (among the economic labor force) |  |  |  |  |  |  |
| Yes | 450 | 95.3 | (93.4, 97.2) | 220 | 72.6 | (67.6, 77.7) |
| No | 22 | 4.7 | (2.8, 6.6) | 83 | 28.4 | (22.3, 32.4) |
| Health insurance coverage over the last 12 months |  |  |  |  |  |  |
| 0 | 12 | 1.7 | (0.7, 2.6) | 55 | 12.3 | (9.3, 15.4) |
| 1 to 11 | 30 | 4.1 | (2.7, 5.6) | 50 | 11.2 | (8.3, 14.2) |
| 12 | 681 | 94.2 | (92.5, 95.9) | 341 | 76.5 | (72.5, 80.4) |
| Census 2010 urban / rural classification |  |  |  |  |  |  |
| Urban | 575 | 79.3 | (76.4, 82.3) | 517 | 100.0 | NA |
| Rural | 150 | 20.7 | (17.7, 23.6) | NA | NA | NA |

Frequencies may not add to the total sample size due to missing values.

**Table 2-C.** SHOW Children in WAVES II, III and IV Characteristics.

| Demographic characteristics | WAVE II<br>2014-2016 |  |  | WAVE III<br>2017 |  |  | WAVE IV<br>2018-2019 |  |  |
| --- | --- | --- | --- | --- | --- | --- | --- | --- | --- |
|  | N* | Mean or<br>%** | Range<br>95% CI** or | N* | Mean or<br>%* | Range<br>95% CI* or | N* | Mean or<br>%* | Range or<br>95% CI* |
| Age (years) | 645 | 7.7 | 0 - 17 | 222 | 8.6 | 0 - 17 | 113 | 8.0 | 0 - 17 |
| 0-6 | 279 | 44.8 | (39.4, 50.2) | 71 | 32.0 | (25.8, 38.2) | 49 | 43.4 | (34.1, 52.6) |
| 7-11 | 182 | 28.1 | (25.5, 30.8) | 88 | 39.6 | (33.2, 46.1) | 27 | 23.9 | (15.9, 31.9) |
| 12-17 | 184 | 27.1 | (22.2, 31.9) | 63 | 28.4 | (22.4, 34.4) | 37 | 32.7 | (24.0, 41.5) |
| Gender |  |  |  |  |  |  |  |  |  |
| Male | 332 | 51.1 | (46.5, 55.8) | 123 | 55.4 | (48.8, 62.0) | 59 | 52.2 | (42.9, 61.6) |
| Female | 313 | 48.9 | (44.2, 53.5) | 99 | 44.6 | (38.0, 51.2) | 54 | 47.8 | (38.4, 57.2) |
| Race / ethnicity |  |  |  |  |  |  |  |  |  |
| Non-Hispanic white | 472 | 71.4 | (62.9, 79.8) | 149 | 67.4 | (61.2, 73.6) | 4 | 3.6 | (0.1, 7.1) |
| Non-Hispanic black | 103 | 16.8 | (8.2, 25.5) | 38 | 17.2 | (12.2, 22.2) | 101 | 90.2 | (84.6, 95.8) |
| Hispanic | 15 | 2.5 | (0.2, 4.7) | 23 | 10.4 | (6.3, 14.5) | 6 | 5.4 | (1.1, 9.6) |
| Other | 53 | 9.3 | (5.5, 13.2) | 11 | 5.0 | (2.1, 7.9) | 1 | 0.9 | (0.0, 2.7) |

\* Unweighted
\*\* Weighted and adjusted for the stratification and clustering in the complex survey sampling design.
Frequencies may not add to the total sample size due to missing values.

Alternative convenience based recruitment strategies were developed collaboratively with and in response to community partners interests in using an asset-based, community-driven model to guide research in the City of Milwaukee. Collaborations were led by investigators with the University of Wisconsin Center for Community Engagement and Health Partnerships (CCEHP).^7^ The partnerships and stakeholders informed all aspects of recruitment, including promotion opportunities, use of community events and modifications to survey content relevant to stakeholder interests. Survey elements were modified for use in Hispanic populations and Spanish translation of the final survey content approved by CCEHP partner organizations. Survey weights are not available for WAVE IV due to the hybrid nature of the sampling approach.

### What has been measured?

Table 3 outlines the breadth of questionnaire, physical exam and biomarker data collected among SHOW participants. SHOW was not originally designed with a specific set of hypotheses in mind but with a broader mission to improve understanding of the multi-level determinants of health and equity, originally emphasizing chronic diseases in adult populations. Thus, the protocols are flexible enough to add new collection tools relevant to study hypotheses as needed.^8^ Tables 4A and 4B describe key findings on health status for WAVES I and II and WAVES II and IV respectively, Supplemental Table 2 highlights the distribution of questionnaires by survey wave.

**Table 3.**
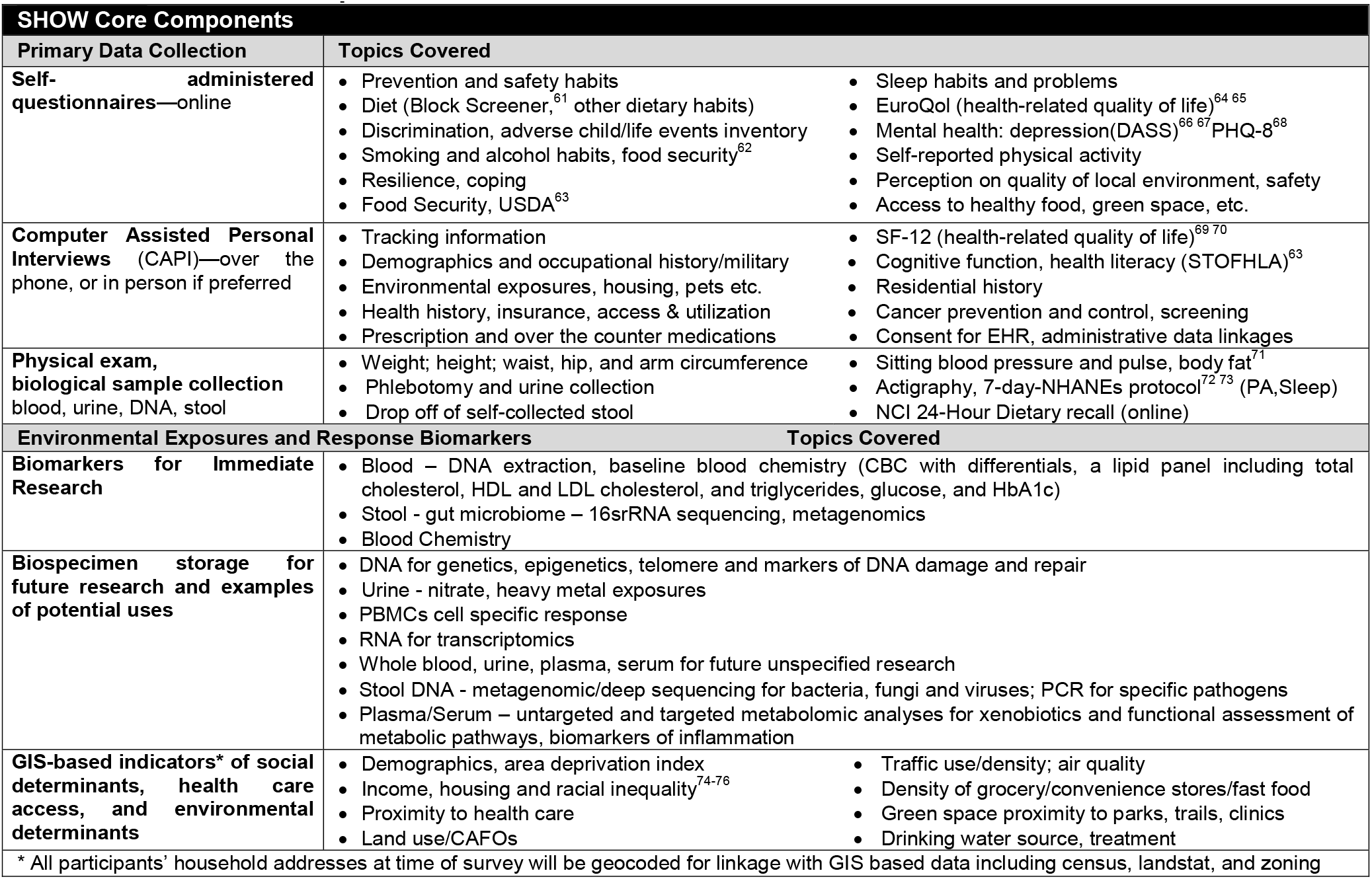
SHOW Core Components.

| SHOW Core Components |  |  |
| --- | --- | --- |
| Primary Data Collection | Topics Covered |  |
| Self-administered questionnaires—online | • Prevention and safety habits<br>• Diet (Block Screener,<sup>61</sup> other dietary habits)<br>• Discrimination, adverse child/life events inventory<br>• Smoking and alcohol habits, food security<sup>62</sup><br>• Resilience, coping<br>• Food Security, USDA<sup>63</sup> | • Sleep habits and problems<br>• EuroQol (health-related quality of life)<sup>64 65</sup><br>• Mental health: depression(DASS)<sup>66 67</sup> PHQ-8<sup>68</sup><br>• Self-reported physical activity<br>• Perception on quality of local environment, safety<br>• Access to healthy food, green space, etc. |
| Computer Assisted Personal Interviews (CAPI)—over the phone, or in person if preferred | • Tracking information<br>• Demographics and occupational history/military<br>• Environmental exposures, housing, pets etc.<br>• Health history, insurance, access & utilization<br>• Prescription and over the counter medications | • SF-12 (health-related quality of life)<sup>69 70</sup><br>• Cognitive function, health literacy (STOFHLA)<sup>63</sup><br>• Residential history<br>• Cancer prevention and control, screening<br>• Consent for EHR, administrative data linkages |
| Physical exam, biological sample collection<br>blood, urine, DNA, stool | • Weight; height; waist, hip, and arm circumference<br>• Phlebotomy and urine collection<br>• Drop off of self-collected stool | • Sitting blood pressure and pulse, body fat<sup>71</sup><br>• Actigraphy, 7-day-NHANEs protocol<sup>72 73</sup> (PA, Sleep)<br>• NCI 24-Hour Dietary recall (online) |
| Environmental Exposures and Response Biomarkers |  |  |
| Biomarkers for Immediate Research | • Blood – DNA extraction, baseline blood chemistry (CBC with differentials, a lipid panel including total cholesterol, HDL and LDL cholesterol, and triglycerides, glucose, and HbA1c)<br>• Stool - gut microbiome – 16srRNA sequencing, metagenomics<br>• Blood Chemistry |  |
| Biospecimen storage for future research and examples of potential uses | • DNA for genetics, epigenetics, telomere and markers of DNA damage and repair<br>• Urine - nitrate, heavy metal exposures<br>• PBMCs cell specific response<br>• RNA for transcriptomics<br>• Whole blood, urine, plasma, serum for future unspecified research<br>• Stool DNA - metagenomic/deep sequencing for bacteria, fungi and viruses; PCR for specific pathogens<br>• Plasma/Serum – untargeted and targeted metabolomic analyses for xenobiotics and functional assessment of metabolic pathways, biomarkers of inflammation |  |
| GIS-based indicators* of social determinants, health care access, and environmental determinants | • Demographics, area deprivation index<br>• Income, housing and racial inequality<sup>74-76</sup><br>• Proximity to health care<br>• Land use/CAFOs | • Traffic use/density; air quality<br>• Density of grocery/convenience stores/fast food<br>• Green space proximity to parks, trails, clinics<br>• Drinking water source, treatment |
| * All participants' household addresses at time of survey will be geocoded for linkage with GIS based data including census, landstat, and zoning |  |  |

**Table 4-A.**
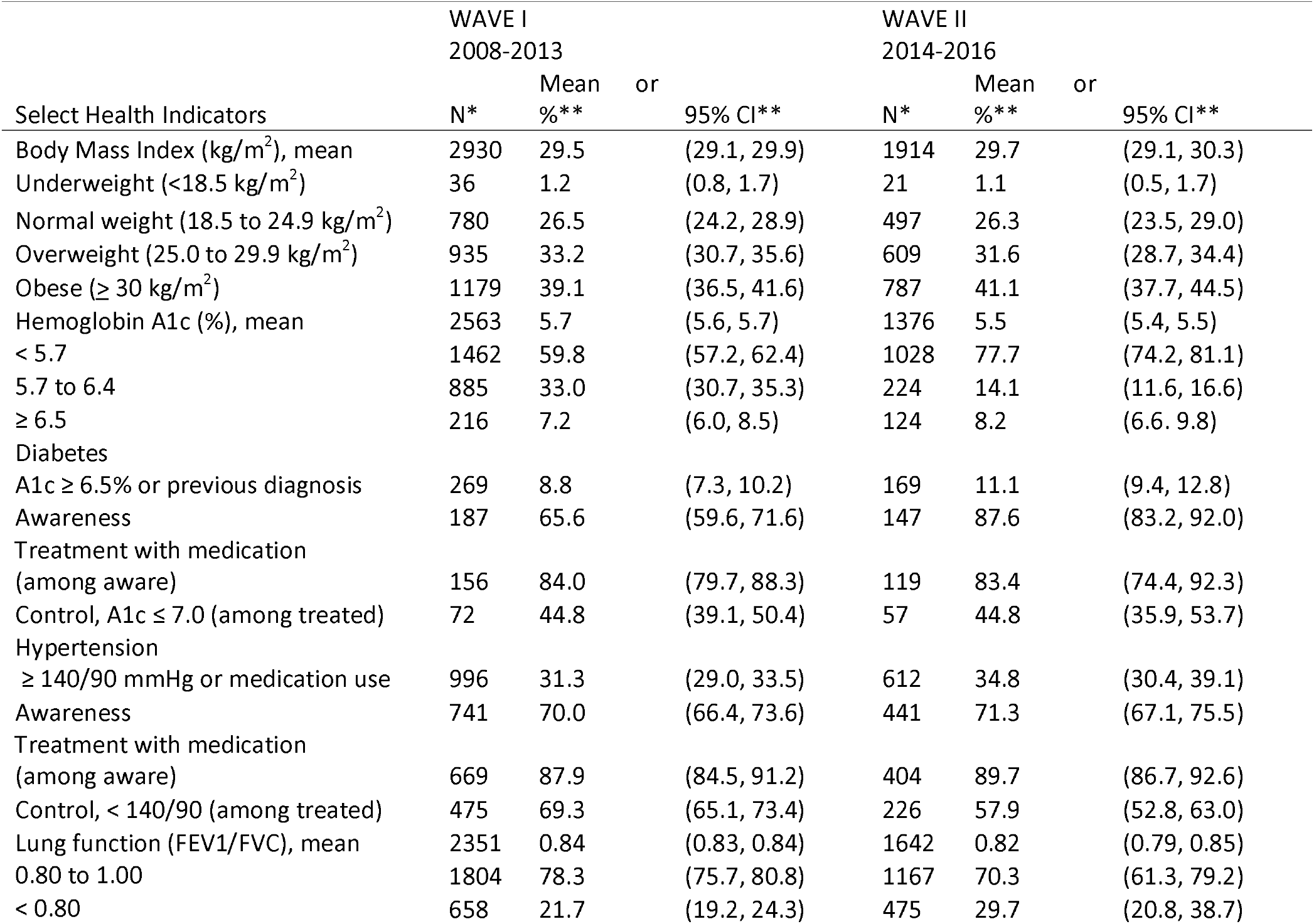
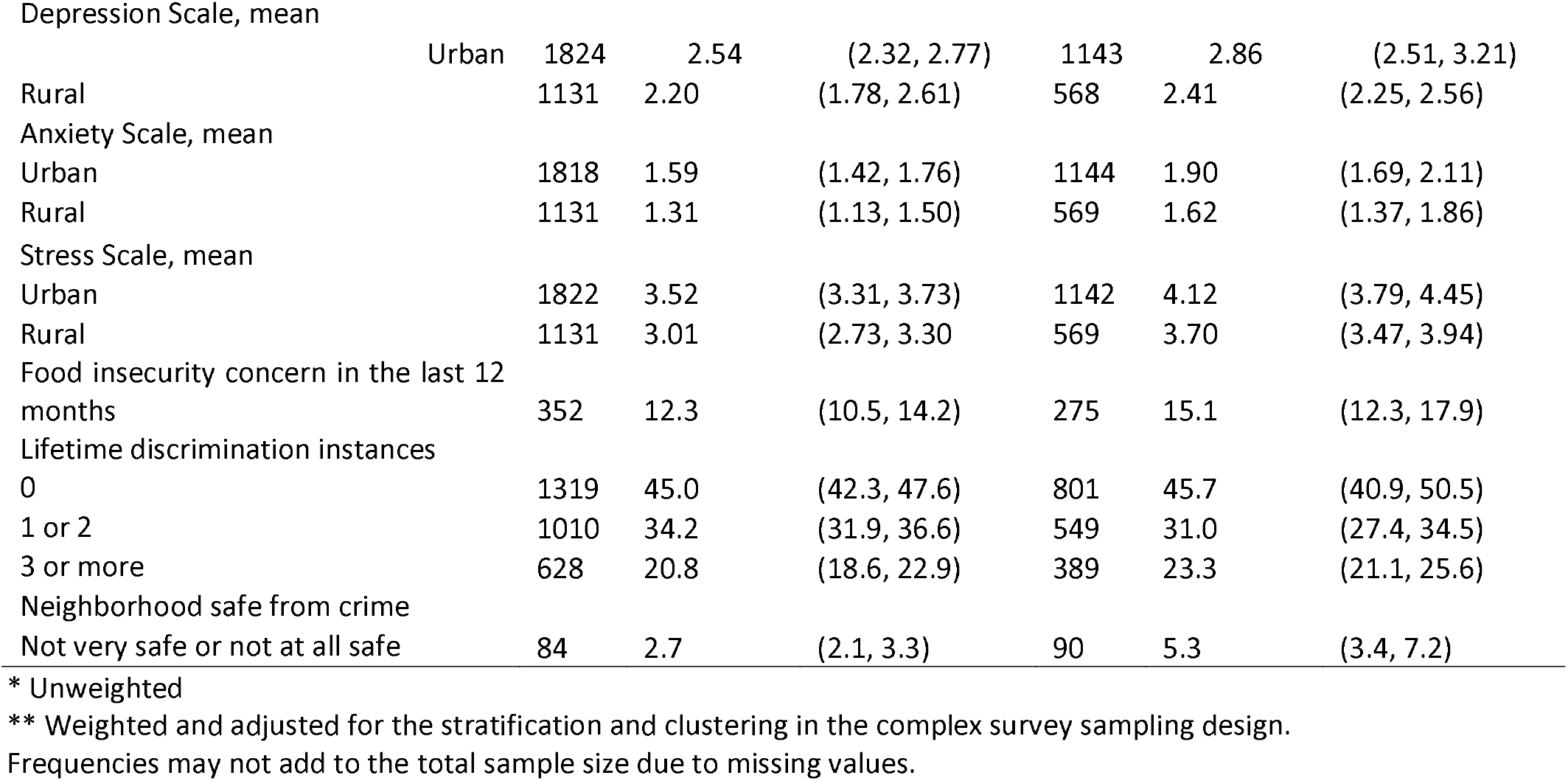
Select Health Indicators for SHOW Adults WAVES I and II, Weighted for Statewide Sample Estimation.

| Select Health Indicators | WAVE I<br>2008-2013 |  |  | WAVE II<br>2014-2016 |  |  |
| --- | --- | --- | --- | --- | --- | --- |
|  | N* | Mean<br>%** | or<br>95% CI** | N* | Mean<br>%** | or<br>95% CI** |
| Body Mass Index (kg/m<sup>2</sup>), mean | 2930 | 29.5 | (29.1, 29.9) | 1914 | 29.7 | (29.1, 30.3) |
| Underweight (<18.5 kg/m<sup>2</sup>) | 36 | 1.2 | (0.8, 1.7) | 21 | 1.1 | (0.5, 1.7) |
| Normal weight (18.5 to 24.9 kg/m<sup>2</sup>) | 780 | 26.5 | (24.2, 28.9) | 497 | 26.3 | (23.5, 29.0) |
| Overweight (25.0 to 29.9 kg/m<sup>2</sup>) | 935 | 33.2 | (30.7, 35.6) | 609 | 31.6 | (28.7, 34.4) |
| Obese (≥ 30 kg/m<sup>2</sup>) | 1179 | 39.1 | (36.5, 41.6) | 787 | 41.1 | (37.7, 44.5) |
| Hemoglobin A1c (%), mean | 2563 | 5.7 | (5.6, 5.7) | 1376 | 5.5 | (5.4, 5.5) |
| < 5.7 | 1462 | 59.8 | (57.2, 62.4) | 1028 | 77.7 | (74.2, 81.1) |
| 5.7 to 6.4 | 885 | 33.0 | (30.7, 35.3) | 224 | 14.1 | (11.6, 16.6) |
| ≥ 6.5 | 216 | 7.2 | (6.0, 8.5) | 124 | 8.2 | (6.6, 9.8) |
| Diabetes |  |  |  |  |  |  |
| A1c ≥ 6.5% or previous diagnosis | 269 | 8.8 | (7.3, 10.2) | 169 | 11.1 | (9.4, 12.8) |
| Awareness | 187 | 65.6 | (59.6, 71.6) | 147 | 87.6 | (83.2, 92.0) |
| Treatment with medication |  |  |  |  |  |  |
| (among aware) | 156 | 84.0 | (79.7, 88.3) | 119 | 83.4 | (74.4, 92.3) |
| Control, A1c ≤ 7.0 (among treated) | 72 | 44.8 | (39.1, 50.4) | 57 | 44.8 | (35.9, 53.7) |
| Hypertension |  |  |  |  |  |  |
| ≥ 140/90 mmHg or medication use | 996 | 31.3 | (29.0, 33.5) | 612 | 34.8 | (30.4, 39.1) |
| Awareness | 741 | 70.0 | (66.4, 73.6) | 441 | 71.3 | (67.1, 75.5) |
| Treatment with medication |  |  |  |  |  |  |
| (among aware) | 669 | 87.9 | (84.5, 91.2) | 404 | 89.7 | (86.7, 92.6) |
| Control, < 140/90 (among treated) | 475 | 69.3 | (65.1, 73.4) | 226 | 57.9 | (52.8, 63.0) |
| Lung function (FEV1/FVC), mean | 2351 | 0.84 | (0.83, 0.84) | 1642 | 0.82 | (0.79, 0.85) |
| 0.80 to 1.00 | 1804 | 78.3 | (75.7, 80.8) | 1167 | 70.3 | (61.3, 79.2) |
| < 0.80 | 658 | 21.7 | (19.2, 24.3) | 475 | 29.7 | (20.8, 38.7) |
| Depression Scale, mean |  |  |  |  |  |  |
| Urban | 1824 | 2.54 | (2.32, 2.77) | 1143 | 2.86 | (2.51, 3.21) |
| Rural | 1131 | 2.20 | (1.78, 2.61) | 568 | 2.41 | (2.25, 2.56) |
| Anxiety Scale, mean |  |  |  |  |  |  |
| Urban | 1818 | 1.59 | (1.42, 1.76) | 1144 | 1.90 | (1.69, 2.11) |
| Rural | 1131 | 1.31 | (1.13, 1.50) | 569 | 1.62 | (1.37, 1.86) |
| Stress Scale, mean |  |  |  |  |  |  |
| Urban | 1822 | 3.52 | (3.31, 3.73) | 1142 | 4.12 | (3.79, 4.45) |
| Rural | 1131 | 3.01 | (2.73, 3.30) | 569 | 3.70 | (3.47, 3.94) |
| Food insecurity concern in the last 12 months |  |  |  |  |  |  |
|  | 352 | 12.3 | (10.5, 14.2) | 275 | 15.1 | (12.3, 17.9) |
| Lifetime discrimination instances |  |  |  |  |  |  |
| 0 | 1319 | 45.0 | (42.3, 47.6) | 801 | 45.7 | (40.9, 50.5) |
| 1 or 2 | 1010 | 34.2 | (31.9, 36.6) | 549 | 31.0 | (27.4, 34.5) |
| 3 or more | 628 | 20.8 | (18.6, 22.9) | 389 | 23.3 | (21.1, 25.6) |
| Neighborhood safe from crime |  |  |  |  |  |  |
| Not very safe or not at all safe | 84 | 2.7 | (2.1, 3.3) | 90 | 5.3 | (3.4, 7.2) |

\* Unweighted
\*\* Weighted and adjusted for the stratification and clustering in the complex survey sampling design.
Frequencies may not add to the total sample size due to missing values.

**Table 4-B.** Select Health Indicators for SHOW Adults WAVES III and IV, Unweighted.

| Select Health Indicators | WAVE III<br>2017 |  |  | WAVE IV<br>2018-2019 |  |  |
| --- | --- | --- | --- | --- | --- | --- |
|  | N* | Mean<br>%* | or<br>95% CI** | N* | Mean<br>or %** | 95% CI** |
| Body Mass Index (kg/m<sup>2</sup>), mean | 716 | 30.9 | (30.4, 31.5) | 501 | 32.1 | (31.4, 32.8) |
| Underweight (<18.5 kg/m<sup>2</sup>) | 6 | 0.8 | (0.2, 1.5) | 6 | 1.2 | (0.2, 2.2) |
| Normal weight (18.5 to 24.9 kg/m<sup>2</sup>) | 156 | 21.8 | (18.8, 24.8) | 77 | 15.4 | (12.2, 18.5) |
| Overweight (25.0 to 29.9 kg/m<sup>2</sup>) | 204 | 28.5 | (25.2, 31.8) | 139 | 27.7 | (23.8, 31.7) |
| Obese (≥ 30 kg/m<sup>2</sup>) | 350 | 48.9 | (45.2, 52.6) | 279 | 55.7 | (51.3, 60.1) |
| Hemoglobin A1c (%), mean | 508 | 5.7 | (5.6, 5.8) | 343 | 6.1 | (6.0, 6.3) |
| < 5.7 | 348 | 68.5 | (64.5, 72.6) | 144 | 42.0 | (36.7, 47.2) |
| 5.7 to 6.4 | 114 | 22.4 | (18.8, 26.1) | 129 | 37.6 | (32.4, 42.8) |
| ≥ 6.5 | 46 | 9.1 | (6.6, 11.6) | 70 | 20.4 | (16.1, 24.7) |
| Diabetes |  |  |  |  |  |  |
| A1c ≥ 6.5% or previous diagnosis | 60 | 11.8 | (8.8, 14.8) | 79 | 23.1 | (18.6, 27.6) |
| Awareness | 49 | 81.7 | (71.6, 91.7) | 51 | 65.4 | (54.6, 76.1) |
| Treatment with medication |  |  |  |  |  |  |
| (among aware) | 41 | 83.7 | (72.9, 94.4) | 45 | 88.2 | (79.1, 97.4) |
| Control, A1c ≤ 7.0 (among treated) | 20 | 48.8 | (32.8, 64.8) | 17 | 37.8 | (23.0, 52.5) |
| Hypertension |  |  |  |  |  |  |
| ≥ 140/90 mmHg or medication use | 303 | 42.4 | (38.8, 46.1) | 237 | 48.6 | (44.1, 53.0) |
| Awareness | 223 | 73.6 | (68.6, 78.6) | 207 | 87.3 | (83.1, 91.6) |
| Treatment with medication |  |  |  |  |  |  |
| (among aware) | 200 | 89.7 | (85.7, 93.7) | 188 | 90.8 | (86.9, 94.8) |
| Control, < 140/90 (among treated) | 113 | 56.5 | (49.6, 63.4) | 112 | 59.6 | (52.5, 66.7) |
| Lung function (FEV1/FVC), mean | 652 | 0.84 | (0.83, 0.85) | 292 | 0.81 | (0.79, 0.82) |
| 0.80 to 1.00 | 524 | 80.4 | (77.3, 83.4) | 227 | 61.5 | (56.5, 66.5) |
| < 0.80 | 128 | 19.6 | (16.6, 22.7) | 142 | 38.5 | (33.5, 43.7) |
| Depression Scale, mean |  |  |  |  |  |  |
| Urban | 480 | 2.71 | (2.35, 3.07) | 346 | 4.36 | (3.81, 4.90) |
| Rural | 137 | 2.34 | (1.77, 2.92) | NA | NA | NA |
| Anxiety Scale, mean |  |  |  |  |  |  |
| Urban | 480 | 1.88 | (1.63, 2.12) | 352 | 3.56 | (3.11, 4.02) |
| Rural | 138 | 1.34 | (0.98, 1.70) | NA | NA | NA |
| Stress Scale, mean |  |  |  |  |  |  |
| Urban | 481 | 3.88 | (3.55, 4.21) | 349 | 4.81 | (4.31, 5.32) |
| Rural | 138 | 3.28 | (2.70, 3.85) | NA | NA | NA |
| Food insecurity concern in the last 12 months | 84 | 11.7 | (9.3, 14.0) | 146 | 30.2 | (26.1, 34.3) |
| Lifetime discrimination instances |  |  |  |  |  |  |
| 0 | 287 | 45.7 | (41.7, 49.5) | 88 | 26.0 | (21.3, 30.6) |
| 1 or 2 | 194 | 30.8 | (27.2, 34.5) | 101 | 29.8 | (24.9, 34.7) |
| 3 or more | 148 | 23.5 | (20.2, 26.9) | 150 | 44.2 | (38.9, 49.6) |
| Neighborhood safe from crime |  |  |  |  |  |  |
| Not very safe or not at all safe | 39 | 6.2 | (4.3, 8.1) | 141 | 38.3 | (33.3, 43.3) |

Frequencies may not add to the total sample size due to missing values.

#### Interviews and questionnaires

The in-home visit by field interviewers includes computer-assisted personal interviews (CAPI) to gather information on health history and important covariates such as occupation, home environment, health care access, medication use, and demographics.^4^ Several self-administered questionnaires either on paper or increasingly offered online are used to gather detailed information capturing a broad array of social determinants including food security and economic hardship, personal and family medical history, mental health and well-being, quality of life, every day and lifetime racial and other discrimination, life evets, resilience, and coping scales. A neighborhood perceptions questionnaire captures community assets and perceived neighborhood stressors. A personal exposure history^9-11^, includes information on residential history, household characteristics including the age of the home, pet ownership, use of indoor/outdoor pesticides, and smoking policies and water source (private well vs. municipal) ^12^ including use of water filtration. Health behaviors include physical activity, diet, sleep, smoking, and drug and alcohol use. Usual and most recent diet information are captured using both the NCI food frequency questionnaire (all WAVES) and the 24-hour dietary recall.

#### Physical and clinical measurements

In addition to survey data, participants undergo a brief physical exam that includes standardized measurements of blood pressure, weight, height, waist and hip circumference, respiratory function, and collection of blood and urine samples. Weight is measured in kilograms (to a precision of ±0.1 kg) using digital scales with subjects wearing light clothing or surgical scrubs. Height, hip and waist circumference (all in cm) are measured twice. Sitting blood pressure and heart rate are measured using digital blood monitors with three measurements taken one minute apart after an initial 5-minute rest period. Lung function is assessed by spirometry using a Jaeger AM1+ electronic peak flow meter with filter mouthpiece. Testing provides data on FEV1 (forced expiratory volume in 1 second) and FVC (forced vital capacity).

#### Wearable Measurement of Objective physical activity and sleep measurements

Objective physical activity and sleep data are obtained using wearable technology. A detailed protocol for participant 7-day hip and wrist protocol using ActiGraph wGT3X-BT accelerometers (ActiGraph, Pensacola, FL) was developed for both adults, and children >6 years. Data are processed and analyzed using ActiLife software. Both raw and processed data are made available to investigators.

#### Biosample collection and biobanking

All participants providing biological samples are also asked to consent for use of these biological samples for DNA analyses and other future unspecified research. The growing biobank includes over 200,000 cryovials of urine, plasma, serum, PaxGene and DNA samples stored at -80 C for future unspecified research. Following an in-home visit, biological samples are collected either in participant homes or at local exam centers. Several tubes of venous blood (about 55-60 ml in total) are collected and immediately processed for serum and plasma, aliquoted into cryovials and frozen at -80C. A blood aliquot is sent to Marshfield Labs (Marshfield, WI) for complete blood cell count with differential, hematocrit, hemoglobin, HbA1c, glucose, creatinine, triglycerides, total and HDL cholesterol. Blood samples are sent to Prevention Genetics (Marshfield, WI) for DNA extraction. Urine samples are centrifuged, aliquoted into cryovials and frozen. Starting in WAVE II, PAXgene tubes for RNA extraction were added to the collection protocol.

In 2016, ancillary study funding supported expansion of biological sample collection to stool, nasal, and skin swabs for microbiome analyses. Stool specimens are self-collected using a commercial “toilet hat” collection kit within 12 hours of the exam visit. Our current studies have over 95% adherence to this self-collection protocol, including shipping specimens in the correct containers and temperature.

DNA from a subset of n=650 participants were analyzed by the NIH Center for Inherited Disease Research (CIDR). The program provided genome-wide MEGA chip array data for identification of SNP polymorphisms, and DNA methylation for epigenetic analyses. The same subset of 650 individuals also have stool microbiome data available.

#### Linkages with Extant Environmental and Socio-Demographic Data

All participants are geocoded to the household address level that can linked to social and environmental data at multiple geographic scales. In addition, all participants are consented for linkage with administrative databases including vital statistics and state cancer registry data. Ongoing efforts are being made to reconsent participants for linkage with electronic medical records and for deposition of genetic and epigenetic analyses into NIH dbGaP database. Socio-demographic and environmental measures can be linked to the data using a street address or other geography indicators (e.g., CBG). Environmental measures include air pollution exposure (fine particulate matter and traffic pollution), ^13 14^ access to retail food outlets,^15^ access to health care facilities,^16^ measures of green space (vegetation index via satellite imagery and percent coverage from a tree canopy database)^17 18^ and drinking water source.^19^

### Ancillary Studies

Numerous ancillary studies have either extended the focus of the baseline SHOW program or facilitated follow-up with cohort participants around particular etiologic, prevention or intervention research questions. Examples include personalized vitamin D supplementation based on genetic analysis,^20^ impacts of caregiver strain on telomere length and quality of life,^21-23^ assessment of physical activity in rural women,^24 25^ incontinence research in older women,^26^ examining how household context impacts personal health information management,^27-29^ chronic stress and cardio-metabolic risk,^14 30^ and epigenetic signatures of aging and health disparities, among others. SHOW also supports applied public health and surveillance. Examples of projects with the Wisconsin Department of Health include oral health screening,^31 32^ as well as a long-standing collaboration to examine the health impacts of Great Lakes fish consumption across the state, among anglers and in high-risk populations (e.g., Burmese immigrants).^33-38^

#### Wisconsin Microbiome and Other NIH Funded Research

In 2016, The Wisconsin Microbiome Study, was launched to investigate the presence of multi-drug resistant organisms (MDROs) and to characterize the human microbiome in the population.^39^ SHOW added questionnaires on risk factors for MDRO colonization, diet history, and food-frequency. Stool and swab samples (skin, nasal, oral) were collected from 700 participants and analyzed for MDRO colonization; 16s rRNA gene sequencing data are available for all stool samples collected with this project. In 2018, 50% of Wisconsin Microbiome Study participants were invited to complete a follow-up visit. Stool and environmental samples (high-touch surface swab, household dust, and soil samples) were collected and are available for future analyses. Additional NIH research funded by the National Institutes of Aging and The National Institute of Allergy and Infectious diseases are ongoing.

### Key Findings to Date

The breadth and nature of data collected by the SHOW program allows for multidisciplinary research on various health topics. The main findings to-date have focused on population health priorities including obesity, cardiometabolic and pulmonary health, mental health, and cancer prevention and control.^13-17 32 40-44^ SHOW supports comprehensive assessment of health disparities, associated with neighborhood environment, access to healthy food, health care, oral health and experiences of discrimination.^14-16^ Food insecurity is highly prevalent in inner city and rural communities across the state, with several adverse metabolic and cardiovascular outcomes.^42 45^ SHOW has also supported research on biological effects of multiple social determinants of health including caregiver strain,^21-23^ and neighborhood stress.^14^ Objective and subjective measures of physical activity and the built environment continue to support novel methods for behavioral and built environment research in both child and adult populations.^24 46-49^ The complete list of over 60 publications is available at www.med.wisc.edu/show. Below is a brief summary of key findings including those related to COVID-19 follow-up.

### COVID-19 Impacts on Population Health

Recently, a subset of SHOW study population participated in two COVID-19 specific research efforts which are described in more detail elsewhere.^50 51^ In brief, the randomly selected population-based study has provided a robust platform for COVID-19 antibody surveillance in collaboration with the Wisconsin Department of Health Services and the Wisconsin State Laboratory of Hygiene, the only randomly selected statewide sample to date.^47^ The SHOW program also is currently conducting online survey of COVID-19 impacts on health and well-being over time (May-June, 2020; January-February, 2021; and later May-June 2021) in past SHOW participants. ^46^

#### Environmental Health and Microbiome Research

SHOW was among the first to examine associations between green space and mental health, now a growing area of research.^17^ We found that a positive neighborhood perception and green space correlates with better sleep quality. ^18 52^ Chronic low-level air pollution exposure has shown adverse associations with lung function, and respiratory allergies that vary by level of neighborhood perception of safety and aesthetics. ^13 14^ The Wisconsin Microbiome Ancillary Study in children and adults demonstrated the role of xenobiotics and other settings in shaping the human gut microbiome and increased risk for MDRO colonization.^53-55^ This represents an important and novel area for metabolic, aging and population health research.

#### Obesity and cardiovascular health

Numerous studies examine predictors of obesity, and determinants of metabolic syndrome in the SHOW population.^15 40 42 44 48 56^ Objective measures of obesity indicate that over 70% of the state population is overweight or obese, and that a higher level of obesity is correlated with multiple co-morbidities.^44^ Obesity has also been shown to modify associations of respiratory outcomes with air pollution and smoking exposure in the study sample, suggesting SHOW is a valuable resource for examining the role of obesity in increasing human susceptibility to environmental exposures and the biological mechanisms underlying these associations.

#### Multi-omics Research

Recent analysis of whole blood mRNA levels among SHOW participants revealed differential gene expression in stress and toxicity pathways in obese smokers compared to non-obese smokers ^57^. This work highlights the potential for SHOW to serve as an infrastructure for emerging precision-health initiatives. In 2018, the MEGA Chip Array and EPIC Chip Array analysis was performed on a subset of Wave II SHOW participants that will enable future investigations of gene-environment interactions and studies of biological pathway mediating effects of social determinants on health including epigenetics modifications via DNA methylation pathways.

#### Community and policy research

The program also offers opportunities for measuring the impact of natural experiments related to significant policy changes.^41^ For example, a survey of private well-owners in rural communities found limited knowledge, education, and resources to be barriers to well testing, a known evidence-based strategy for identifying potential adverse environmental exposures in drinking water supplies.^19^ Examples of community-based research include use of abbreviated SHOW surveys in the community to promote community-driven health assessments,^58^ the implementation of an “eating smart” intervention to promote healthy eating,^59 60^ and the objective assessment of the social and built environment.^48^

## Further Details

### Strengths

SHOW was designed using rigorous sampling strategies and provides high quality measures of health and well-being that are comparable to other well-known surveys including the National Health and Nutrition Exam Survey. A breadth of objective and subjective data (over 2000 variables) from a diverse statewide sample offer an invaluable resource for population health research. The biosamples support rigorous translational research including novel biomarkers of response to environmental exposures. Availability of DNA and RNA provides opportunities for future precision health and omics-integration (genomic, epigenomic and transcriptomic) projects. Similarly, stool, plasma/serum and urine samples offer new opportunities for metabolomics and exposure assessment. The program serves as a cost-effective research infrastructure allowing for investigator-initiated ancillary studies. Existing baseline data support future interventions and community-based partnerships for program planning and evaluation. Major strengths of the program also include the ability to link SHOW data to other databases and registers including vital statistics, state cancer registry, and environmental exposure data.

The SHOW program also offers an opportunity to study aging across the life course, including a well-characterized large young-adult, middle-aged, and older adult population. Middle-aged adulthood is a time when many pathological changes of disorders begin, but are still clinically undetectable. Thus, SHOW population samples enable studies exploring early biomarkers of age-related disorders and the potential for long-term follow-up. Increasingly new models of research are looking toward electronic health records for understanding health trajectories over time. SHOW also has consented individuals for linkages with electronic records and other administrative data, allowing for new efforts in data integration, and method validation to emerge. Many additional ongoing ancillary studies are capitalizing on this infrastructure for advancing multi-level population health research in children, adults and among under-represented populations. A recent focus of the program has been community engagement and outreach among minority populations and rurally isolated populations to identify opportunities to collect additional data and leverage additional resources to support community-based intervention work.

The SHOW sample includes a significant number of genetically related (parent-child; siblings) and unrelated (husband-wife) participants with similar exposures or lifestyles. Such sample structure allows various types of investigations on health determinants and variability in human responses to similar factors.

### Limitations

Conducting SHOW as a comprehensive population-based survey is both resource- and time-intensive. SHOW’s sampling strategy was designed to ensure a statewide representative sample leading to both logistical and monetary costs. Although the resulting sample characteristics may be a strength for many types of epidemiological studies, it may be a limitation for other studies requiring a more substantial proportion of non-white participants, as the vast majority of state residents are white and less than 12% of the state’s total population self-identifies as non-white. SHOW has recognized this limitation and in 2018-2019 conducted focused recruitment of persons of color in highly diverse communities.

## Data Availability

Data in this article may be obtained by emailing study investigators, or making a formal data request.

https://show.wisc.edu/

## Data Availability

Any qualified researcher, or community academic or applied public health practitioner can request data and biospecimens from the SHOW biobank. A public use data set including sampling weights and use of sampling weights for analyzing SHOW data will be made available on the SHOW website www.show.wisc.edu in the near future. Details on survey instruments and variables and request forms for restricted data (data with unique geographic identifiers, biological samples, genetic and epigenetic, and microbiome data) are also available. SHOW data science core supports students and other faculty in use of SHOW data. SHOW also provides consultation services on the use of SHOW for future ancillary studies, including longitudinal follow-up of select or the full cohort sample.

### Patient and Public Involvement

The core survey contents were determined using a social determinants of health framework to prioritize questions. Whenever possible, questions were selected from previously validated questionnaires. Several ancillary study projects have been done in collaboration with community partners who have extracted a smaller number of survey questions important for goals and dissemination. The core infrastructure values community engagement in all aspects of ancillary study development. Trained field interviewers review consent documents and checklists to assure that participants are informed of all aspects of survey participation prior to consent. Participants may choose to not answer any questions and that they are not required to complete all SHOW components. Incentives for the participation in the program are offered and vary by completion of each survey component. Anonymous feedback forms with self-addressed stamped envelopes are provided to participants following completion of the survey. Participants are allowed to opt out of data sharing for future unspecified research and can opt out of any future participation. SHOW has also obtained an NIH Certificate of Confidentiality, to further ensure data will not be shared for reasons outside the original scope of the survey.

### Funding Declaration

This work was supported by the Wisconsin Partnership Program PERC Award [233 PRJ 25DJ and WPP4444], the National Institutes of Health’s Clinical and Translational Science Award [5UL RR025011] and the National Heart Lung and Blood Institute [1 RC2 HL101468]. Ongoing ancillary study funding from the National Institutes of Health Include [X01HG010110], [R21AI142481] and [R01AG061080].

### Authorship Contributions

KMCM is the program Principal Investigator and is accountable for all aspects of the work and will ensure that all questions related to the accuracy or integrity of any part of the work are appropriately investigated and resolved. All authors contributed to the planning, and conduct of the SHOW cohort including contributions to the design, acquisition or analysis of the work. Authors responsible for drafting this manuscript or revising it critically for important content: KMCM, TJL, MN, AB, CDE, FJN, PEP. Final approval of the version published was made by: KMCM, AB, MN, AS, FJN.

### Conflicts of Interest

Authors have no competing financial or other conflict of interests to declare with this work.

## Acknowledgements

The authors would like to thank the University of Wisconsin Survey Center, SHOW administrative, field, and scientific staff, as well as all the SHOW participants for their contributions to this study.

---

**Supplemental Table 1A.** **SHOW Participation by Health Region or County by Waves I - III, 2008-2017.**

| **Health Region** | **WAVE I**<br>**2008-2013** | **County in**<br>**Health Region** | **WAVE II**<br>**2014-2016** | **WAVE III**<br>**2017** |
| --- | --- | --- | --- | --- |
| North | 66.5% | Wood | 60.1% | 100% |
| Northeast | 57.8% | Brown<br>Waushara | 64.7%<br>80.2% | 82.3% |
| South | 62.0% | Dane | 65.3% | 88.8% |
| Southeast | 50.8% | Milwaukee<br>Racine<br>Ozaukee | 54.4%<br>63.1%<br>63.0% | 85.3% |
| West | 59.9% | La Crosse<br>Eau Claire<br>Polk | 53.0%<br>71.7%<br>73.1% | 80.0% |

**Supplemental Table 1B.**
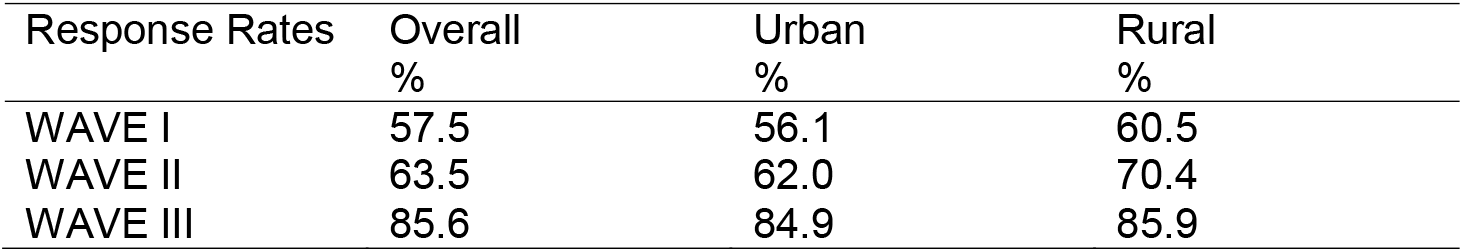
SHOW Participation Rates by Phase and Urbanicity. Rates are estimated as the percent of adult individuals who screened eligible who agree to participate based on cohort year and urban/rural status of resident census tract. A more detailed summary of participation rates by health region (2008-2013 and 2017) and by County (2014-2016) is presented in Supplemental Table 1 and available online.

**Supplemental Table 2:**
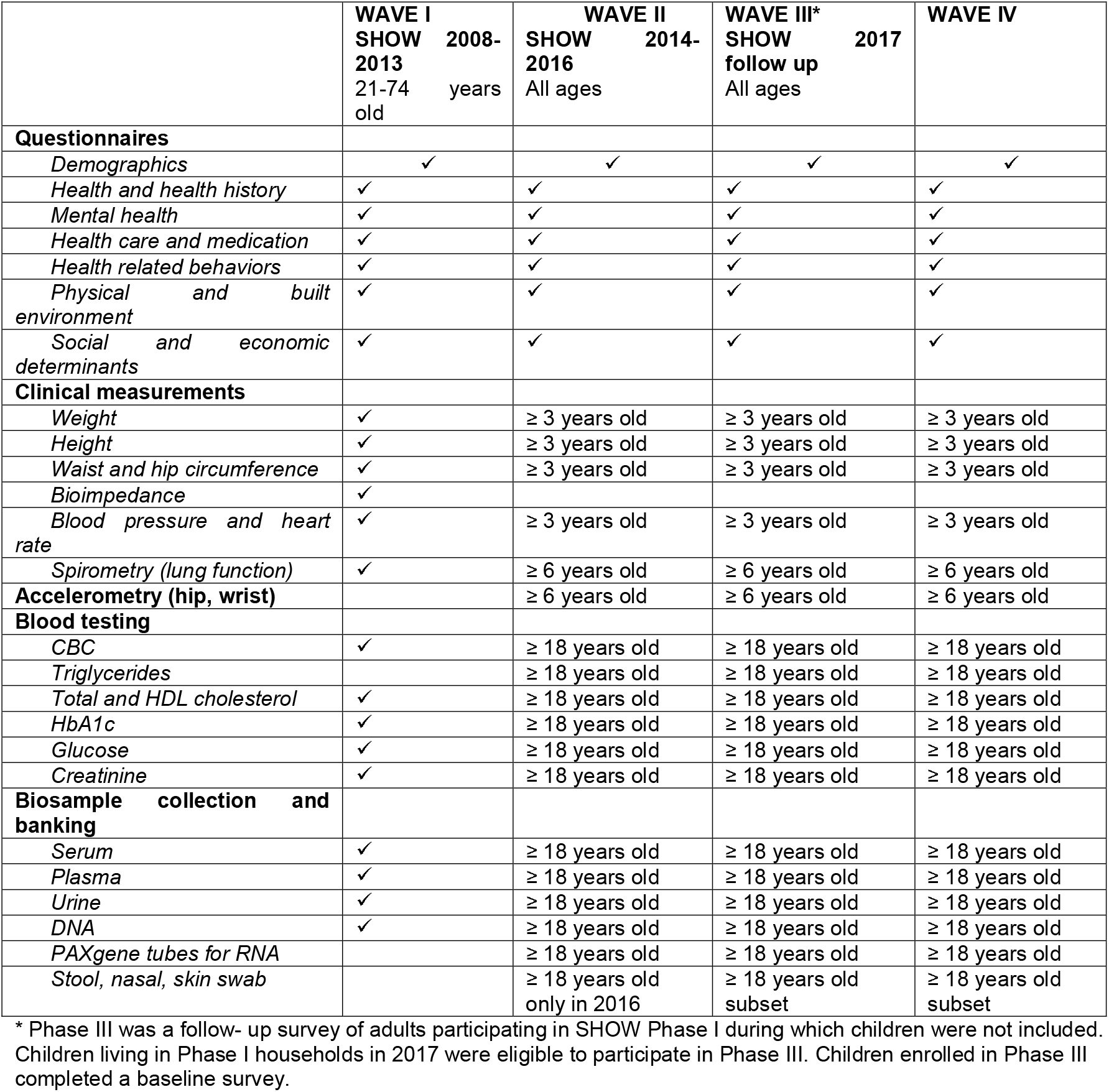
Survey components WAVES I-IV.

**Figure 1.**
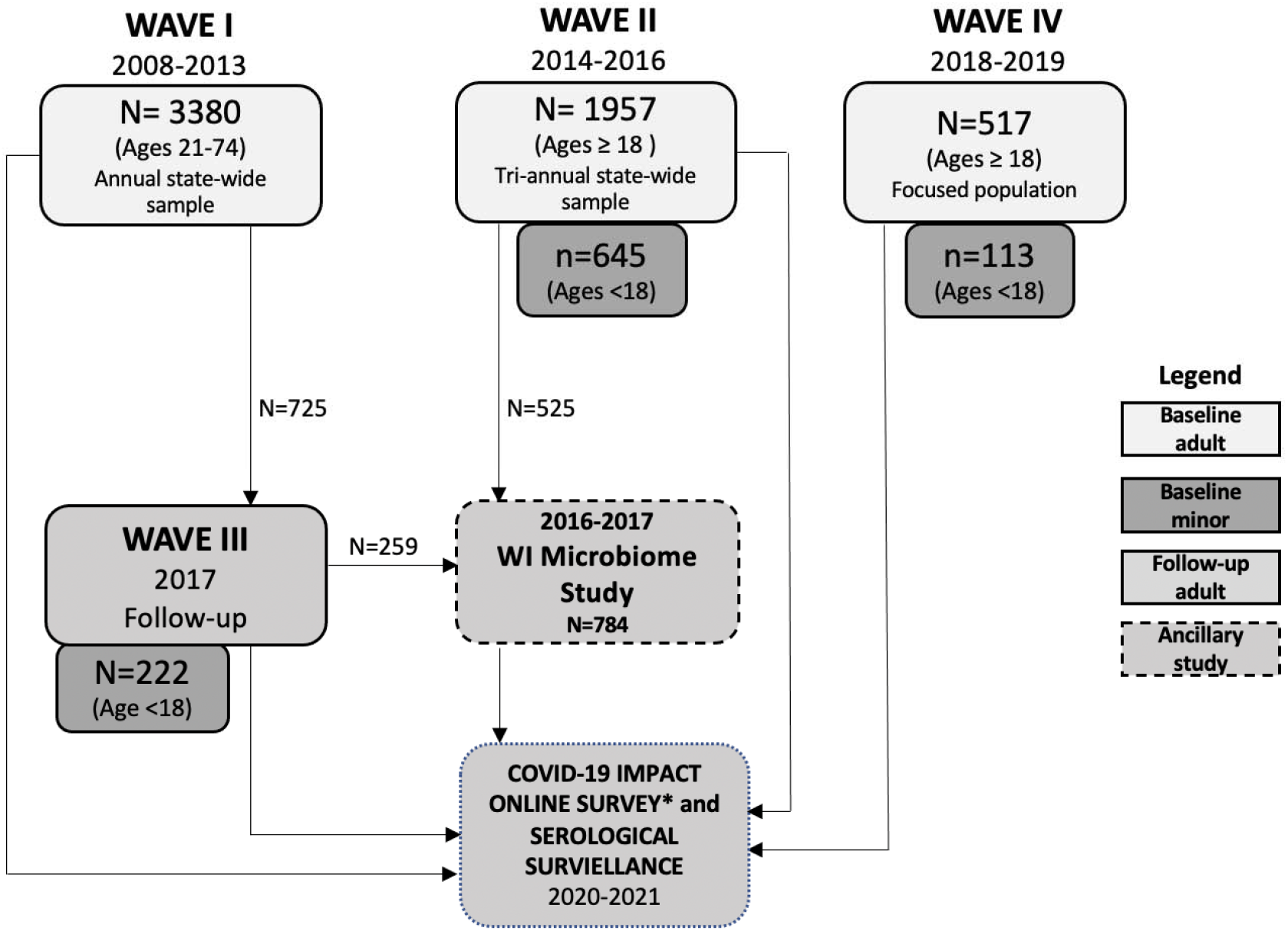
SHOW Survey WAVES and Follow-up Participation (through February 2021)

## Notes

### Competing Interest Statement

The authors have declared no competing interest.

### Clinical Protocols

https://show.wisc.edu/

### Author Declarations

University of Wisconsin Health Sciences Institutional Review Board 2014-0251

### Summary of Updates

This manuscript final figure was revised, abstract revised, and text edited for streamline and clarity.

## REFERENCES

1. Belanger CF, Hennekens CH, Rosner B, et al. The nurses’ health study. Am J Nurs 1978;78(6):1039–40. [published Online First: 1978/06/01]

2. Bao Y, Bertoia ML, Lenart EB, et al. Origin, Methods, and Evolution of the Three Nurses’ Health Studies. American journal of public health 2016;106(9):1573–81. doi: 10.2105/AJPH.2016.303338 [published Online First: 2016/07/26]

3. Tsao CW, Vasan RS. Cohort Profile: The Framingham Heart Study (FHS): overview of milestones in cardiovascular epidemiology. Int J Epidemiol 2015;44(6):1800–13. doi: 10.1093/ije/dyv337

4. Nieto FJ, Peppard PE, Engelman CD, et al. The Survey of the Health of Wisconsin (SHOW), a novel infrastructure for population health research: rationale and methods. BMC Public Health 2010;10:785. doi: 10.1186/1471-2458-10-785 [published Online First: 2010/12/25]

5. Encyclopedia of Survey Research Methods. 2008 doi: 10.4135/9781412963947

6. Nieto FJ, Peppard PE, Engelman CD, et al. The Survey of the Health of Wisconsin (SHOW), a novel infrastructure for population health research: rationale and methods. BMC public health 2010;10(1):785.

7. Green-Harris G, Coley SL, Koscik RL, et al. Addressing Disparities in Alzheimer’s Disease and African-American Participation in Research: An Asset-Based Community Development Approach. Front Aging Neurosci 2019;11:125. doi: 10.3389/fnagi.2019.00125 [published Online First: 2019/06/20]

8. Burke LE, Shiffman S, Music E, et al. Ecological Momentary Assessment in Behavioral Research: Addressing Technological and Human Participant Challenges. J Med Internet Res 2017;19(3):e77–e77. doi: 10.2196/jmir.7138

9. Schultz AA, Malecki KMC, Olson MM, et al. Investigating Cumulative Exposures among 3- to 4-Year-Old Children Using Wearable Ultrafine Particle Sensors and Language Environment Devices: A Pilot and Feasibility Study. Int J Environ Res Public Health 2020;17(14) doi: 10.3390/ijerph17145259 [published Online First: 2020/07/28]

10. Addissie YA, Troia A, Wong ZC, et al. Identifying environmental risk factors and gene-environment interactions in holoprosencephaly. Birth Defects Res 2020 doi: 10.1002/bdr2.1834 [published Online First: 2020/10/29]

11. Addissie YA, Kruszka P, Troia A, et al. Prenatal exposure to pesticides and risk for holoprosencephaly: a case-control study. Environ Health 2020;19(1):65. doi: 10.1186/s12940-020-00611-z [published Online First: 2020/06/10]

12. Malecki KM, Schultz AA, Severtson DJ, et al. Private-well stewardship among a general population based sample of private well-owners. Science of the Total Environment 2017;601:1533–43.

13. Schultz AA, Schauer JJ, Malecki KM. Allergic disease associations with regional and localized estimates of air pollution. Environ Res 2017;155:77–85. doi: 10.1016/j.envres.2017.01.039 [published Online First: 2017/02/15]

14. Malecki KMC, Schultz AA, Bergmans RS. Neighborhood Perceptions and Cumulative Impacts of Low Level Chronic Exposure to Fine Particular Matter (PM2.5) on Cardiopulmonary Health. Int J Environ Res Public Health 2018;15(1) doi: 10.3390/ijerph15010084 [published Online First: 2018/01/11]

15. Laxy M, Malecki KC, Givens ML, et al. The association between neighborhood economic hardship, the retail food environment, fast food intake, and obesity: findings from the Survey of the Health of Wisconsin. BMC Public Health 2015;15:237. doi: 10.1186/s12889-015-1576-x [published Online First: 2015/04/18]

16. Beyer KM, Malecki KM, Hoormann KA, et al. Perceived Neighborhood Quality and Cancer Screening Behavior: Evidence from the Survey of the Health of Wisconsin. J Community Health 2016;41(1):134–7. doi: 10.1007/s10900-015-0078-1 [published Online First: 2015/08/16]

17. Beyer KM, Kaltenbach A, Szabo A, et al. Exposure to neighborhood green space and mental health: evidence from the survey of the health of Wisconsin. Int J Environ Res Public Health 2014;11(3):3453–72. doi: 10.3390/ijerph110303453 [published Online First: 2014/03/26]

18. Johnson BS, Malecki KM, Peppard PE, et al. Exposure to neighborhood green space and sleep: evidence from the Survey of the Health of Wisconsin. Sleep Health 2018;4(5):413–19. doi: 10.1016/j.sleh.2018.08.001 [published Online First: 2018/09/23]

19. Malecki KMC, Schultz AA, Severtson DJ, et al. Private-well stewardship among a general population based sample of private well-owners. Sci Total Environ 2017;601-602:1533–43. doi: 10.1016/j.scitotenv.2017.05.284 [published Online First: 2017/06/14]

20. Engelman CD, Bo R, Zuelsdorff M, et al. Epidemiologic study of the C-3 epimer of 25-hydroxyvitamin D(3) in a population-based sample. Clin Nutr 2014;33(3):421–5. doi: 10.1016/j.clnu.2013.06.005 [published Online First: 2013/07/09]

21. Litzelman K, Skinner HG, Gangnon RE, et al. Role of global stress in the health-related quality of life of caregivers: evidence from the Survey of the Health of Wisconsin. Qual Life Res 2014;23(5):1569–78. doi: 10.1007/s11136-013-0598-z [published Online First: 2013/12/11]

22. Litzelman K, Skinner HG, Gangnon RE, et al. The relationship among caregiving characteristics, caregiver strain, and health-related quality of life: evidence from the Survey of the Health of Wisconsin. Qual Life Res 2015;24(6):1397–406. doi: 10.1007/s11136-014-0874-6 [published Online First: 2014/11/28]

23. Litzelman K, Witt WP, Gangnon RE, et al. Association between informal caregiving and cellular aging in the survey of the health of wisconsin: the role of caregiving characteristics, stress, and strain. Am J Epidemiol 2014;179(11):1340–52. doi: 10.1093/aje/kwu066 [published Online First: 2014/05/02]

24. Gorzelitz JS, Malecki KM, Cadmus-Bertram LA. Awareness of Physical Activity Guidelines Among Rural Women. Am J Prev Med 2020;59(1):143–45. doi: 10.1016/j.amepre.2020.01.022 [published Online First: 2020/06/23]

25. Cadmus-Bertram LA, Gorzelitz JS, Dorn DC, et al. Understanding the physical activity needs and interests of inactive and active rural women: a cross-sectional study of barriers, opportunities, and intervention preferences. J Behav Med 2020;43(4):638–47. doi: 10.1007/s10865-019-00070-z [published Online First: 2019/06/15]

26. Brown HW, Wise ME, LeCaire TJ, et al. Reasons Behind Preferences for Community-Based Continence Promotion. Female Pelvic Med Reconstr Surg 2020;26(7):425–30. doi: 10.1097/SPV.0000000000000806 [published Online First: 2020/03/29]

27. Brennan PF, Ponto K, Casper G, et al. Virtualizing living and working spaces: Proof of concept for a biomedical space-replication methodology. J Biomed Inform 2015;57:53– 61. doi: 10.1016/j.jbi.2015.07.007 [published Online First: 2015/07/15]

28. Casper GR, Brennan PF, Arnott Smith C, et al. Health@Home Moves All About the House! Stud Health Technol Inform 2016;225:173–7. [published Online First: 2016/06/23]

29. Casper GR, Flatley Brennan P, Perreault JO, et al. vizHOME--A context-based home assessment: Preliminary implications for informatics. Stud Health Technol Inform 2015;216:842–6. [published Online First: 2015/08/12]

30. Bautista LE, PK B, MM S, et al. The relationship between chronic stress, hair cortisol and hypertension. International Journal of Cardiology and Hypertension 2019;2(August 2019)

31. Malecki K, Wisk LE, Walsh M, et al. Oral health equity and unmet dental care needs in a population-based sample: findings from the Survey of the Health of Wisconsin. Am J Public Health 2015;105 Suppl 3:S466–74. doi: 10.2105/AJPH.2014.302338 [published Online First: 2015/04/24]

32. VanWormer JJ, Acharya A, Greenlee RT, et al. Oral hygiene and cardiometabolic disease risk in the survey of the health of Wisconsin. Community Dent Oral Epidemiol 2013;41(4):374–84. doi: 10.1111/cdoe.12015 [published Online First: 2012/10/31]

33. Christensen K, Werner M, Malecki K. Serum selenium and lipid levels: Associations observed in the National Health and Nutrition Examination Survey (NHANES) 2011-2012. Environ Res 2015;140:76–84. doi: 10.1016/j.envres.2015.03.020 [published Online First: 2015/04/04]

34. Christensen KY, Thompson BA, Werner M, et al. Levels of nutrients in relation to fish consumption among older male anglers in Wisconsin. Environ Res 2015;142:542–8. doi: 10.1016/j.envres.2015.08.005 [published Online First: 2015/08/22]

35. Christensen KY, Thompson BA, Werner M, et al. Levels of persistent contaminants in relation to fish consumption among older male anglers in Wisconsin. Int J Hyg Environ Health 2016;219(2):184–94. doi: 10.1016/j.ijheh.2015.11.001

36. Raymond MR, Christensen KY, Thompson BA, et al. Associations Between Fish Consumption and Contaminant Biomarkers With Cardiovascular Conditions Among Older Male Anglers in Wisconsin. J Occup Environ Med 2016;58(7):676–82. doi: 10.1097/JOM.0000000000000757 [published Online First: 2016/06/03]

37. Christensen KY, Raymond M, Thompson BA, et al. Perfluoroalkyl substances in older male anglers in Wisconsin. Environ Int 2016;91:312–8. doi: 10.1016/j.envint.2016.03.012 [published Online First: 2016/03/24]

38. Knobeloch L, Imm P, Anderson H. Perfluoroalkyl chemicals in vacuum cleaner dust from 39 Wisconsin homes. Chemosphere 2012;88(7):779–83. doi: 10.1016/j.chemosphere.2012.03.082 [published Online First: 2012/05/01]

39. Eggers S, Malecki KM, Peppard P, et al. Wisconsin microbiome study, a cross-sectional investigation of dietary fibre, microbiome composition and antibiotic-resistant organisms: rationale and methods. BMJ Open 2018;8(3):e019450. doi: 10.1136/bmjopen-2017-019450 [published Online First: 2018/03/29]

40. Givens ML, Malecki KC, Peppard PE, et al. Shiftwork, Sleep Habits, and Metabolic Disparities: Results from the Survey of the Health of Wisconsin. Sleep Health 2015;1(2):115–20. doi: 10.1016/j.sleh.2015.04.014 [published Online First: 2016/02/20]

41. Guzman A, Walsh MC, Smith SS, et al. Evaluating effects of statewide smoking regulations on smoking behaviors among participants in the Survey of the Health of Wisconsin. WMJ 2012;111(4):166-71; quiz 72. [published Online First: 2012/09/14]

42. Saiz AM, Jr., Aul AM, Malecki KM, et al. Food insecurity and cardiovascular health: Findings from a statewide population health survey in Wisconsin. Prev Med 2016;93:1–6. doi: 10.1016/j.ypmed.2016.09.002 [published Online First: 2016/11/05]

43. Shin JI, Bautista LE, Walsh MC, et al. Food insecurity and dyslipidemia in a representative population-based sample in the US. Prev Med 2015;77:186–90. doi: 10.1016/j.ypmed.2015.05.009 [published Online First: 2015/05/27]

44. Eggers S, Remington PL, Ryan K, et al. Obesity Prevalence and Health Consequences: Findings From the Survey of the Health of Wisconsin, 2008-2013. WMJ 2016;115(5):238–44. [published Online First: 2017/11/03]

45. Martinez-Donate AP, Riggall AJ, Meinen AM, et al. Evaluation of a pilot healthy eating intervention in restaurants and food stores of a rural community: a randomized community trial. BMC Public Health 2015;15:136. doi: 10.1186/s12889-015-1469-z [published Online First: 2015/04/18]

46. Bailey EJ, Malecki KC, Engelman CD, et al. Predictors of discordance between perceived and objective neighborhood data. Ann Epidemiol 2014;24(3):214–21. doi: 10.1016/j.annepidem.2013.12.007 [published Online First: 2014/01/29]

47. Gorzelitz J, Peppard PE, Malecki K, et al. Predictors of discordance in self-report versus device-measured physical activity measurement. Ann Epidemiol 2018;28(7):427–31. doi: 10.1016/j.annepidem.2018.03.016 [published Online First: 2018/04/24]

48. Malecki KC, Engelman CD, Peppard PE, et al. The Wisconsin Assessment of the Social and Built Environment (WASABE): a multi-dimensional objective audit instrument for examining neighborhood effects on health. BMC Public Health 2014;14:1165. doi: 10.1186/1471-2458-14-1165 [published Online First: 2014/11/14]

49. Gorzelitz J, Peppard PE, Malecki K, et al. Predictors of discordance in self-report versus device-measured physical activity measurement. Annals of epidemiology 2018;28(7):427–31.

50. Malecki KMC, Schultz AA, Nikodemova M, et al. Statewide Impact of COVID-19 on Social Determinants of Health - A First Look: Findings from the Survey of the Health of Wisconsin. medRxiv 2021:2021.02.18.21252017. doi: 10.1101/2021.02.18.21252017

51. Malecki K, Nikodemova M, Schultz A, et al. Population Changes in Seroprevalence among a Statewide Sample in the United States. medRxiv 2020:2020.12.18.20248479. doi: 10.1101/2020.12.18.20248479

52. Hale L, Hill TD, Friedman E, et al. Perceived neighborhood quality, sleep quality, and health status: evidence from the Survey of the Health of Wisconsin. Soc Sci Med 2013;79:16–22. doi: 10.1016/j.socscimed.2012.07.021 [published Online First: 2012/08/21]

53. Eggers S, Malecki KM, Peppard P, et al. Wisconsin microbiome study, a cross-sectional investigation of dietary fibre, microbiome composition and antibiotic-resistant organisms: rationale and methods. BMJ open 2018;8(3):e019450.

54. Eggers S, Safdar N, Sethi AK, et al. Urinary lead concentration and composition of the adult gut microbiota in a cross-sectional population-based sample. Environment international 2019;133:105122.

55. Kates AE, Jarrett O, Skarlupka JH, et al. Household Pet Ownership and the Microbial Diversity of the Human Gut Microbiota. Front Cell Infect Microbiol 2020;10:73. doi: 10.3389/fcimb.2020.00073 [published Online First: 2020/03/19]

56. Said A, Gagovic V, Malecki K, et al. Primary care practitioners survey of non-alcoholic fatty liver disease. Ann Hepatol 2013;12(5):758–65. [published Online First: 2013/09/11]

57. Nikodemova M, Yee J, Carney PR, et al. Transcriptional differences between smokers and non-smokers and variance by obesity as a risk factor for human sensitivity to environmental exposures. Environ Int 2018;113:249–58. doi: 10.1016/j.envint.2018.02.016 [published Online First: 2018/02/21]

58. Bhutani S, Schoeller DA, Walsh MC, et al. Frequency of Eating Out at Both Fast-Food and Sit-Down Restaurants Was Associated With High Body Mass Index in Non-Large Metropolitan Communities in Midwest. Am J Health Promot 2018;32(1):75–83. doi: 10.1177/0890117116660772 [published Online First: 2016/08/31]

59. Escaron AL, Martinez-Donate AP, Riggall AJ, et al. Developing and Implementing “Waupaca Eating Smart”: A Restaurant and Supermarket Intervention to Promote Healthy Eating Through Changes in the Food Environment. Health Promot Pract 2016;17(2):265–77. doi: 10.1177/1524839915612742

60. Martinez-Donate AP, Espino JV, Meinen A, et al. Neighborhood Disparities in the Restaurant Food Environment. WMJ 2016;115(5):251–8. [published Online First: 2017/11/03]

61. Block G, Gillespie C, Rosenbaum EH, et al. A rapid food screener to assess fat and fruit and vegetable intake. American journal of preventive medicine 2000;18(4):284–8. [published Online First: 2000/05/02]

62. Seligman HK, Laraia BA, Kushel MB. Food insecurity is associated with chronic disease among low-income NHANES participants. J Nutr 2010;140(2):304–10. doi: 10.3945/jn.109.112573 [published Online First: 2009/12/25]

63. Baker DW, Williams MV, Parker RM, et al. Development of a brief test to measure functional health literacy. Patient Educ Couns 1999;38(1):33–42. [published Online First: 2003/10/08]

64. EuroQol--a new facility for the measurement of health-related quality of life. The EuroQol Group. Health Policy 1990;16(3):199–208. [published Online First: 1990/11/05]

65. A Standardized Instrument for Use as a Measure of Health Outcome 2012 [Available from: http://www.euroqol.org/eq-5d/what-is-eq-5d.html accessed July 27, 2012.

66. Lovibond PF, Lovibond SH. The structure of negative emotional states: comparison of the Depression Anxiety Stress Scales (DASS) with the Beck Depression and Anxiety Inventories. Behav Res Ther 1995;33(3):335–43. [published Online First: 1995/03/01]

67. Lovibond SH, Lovibond, P.F. Manual for the Depression Anxiety Stress Scales. 2nd ed. ed. Sydney: Psychology Foundation 1995.

68. Kroenke K, Strine TW, Spitzer RL, et al. The PHQ-8 as a measure of current depression in the general population. J Affect Disord 2009;114(1-3):163–73. doi: 10.1016/j.jad.2008.06.026 [published Online First: 2008/08/30]

69. SF-12v2™ Health Survey 2012 [Available from: http://www.sf-36.org/tools/sf12.shtml.

70. Ware J, Jr., Kosinski M, Keller SD. A 12-Item Short-Form Health Survey: construction of scales and preliminary tests of reliability and validity. Med Care 1996;34(3):220–33. [published Online First: 1996/03/01]

71. Moore VC, Parsons NR, Jaakkola MS, et al. Serial lung function variability using four portable logging meters. J Asthma 2009;46(9):961–6. doi: 10.3109/02770900903229677 [published Online First: 2009/11/13]

72. Matthews CE, Chen KY, Freedson PS, et al. Amount of time spent in sedentary behaviors in the United States, 2003-2004. American journal of epidemiology 2008;167(7):875–81. doi: 10.1093/aje/kwm390 [published Online First: 2008/02/28]

73. Troiano RP, Berrigan D, Dodd KW, et al. Physical activity in the United States measured by accelerometer. Med Sci Sports Exerc 2008;40(1):181–8. doi: 10.1249/mss.0b013e31815a51b3 [published Online First: 2007/12/20]

74. CDC Health Disparities and Inequlities Report - United States, 2011. In: Prevention CfDCa, ed. MMWR. Atlanta, GA: CDC, 2011.

75. Kennedy BP, Kawachi I, Prothrow-Stith D. Income distribution and mortality: cross sectional ecological study of the Robin Hood index in the United States. Bmj 1996;312(7037):1004–7. [published Online First: 1996/04/20]

76. Kondo N, Sembajwe G, Kawachi I, et al. Income inequality, mortality, and self rated health: meta-analysis of multilevel studies. Bmj 2009;339:b4471. doi: 10.1136/bmj.b4471 [published Online First: 2009/11/12]

